# COVID-19 vaccination coverage and hesitancy among Australians with disability and long-term health conditions

**DOI:** 10.1101/2021.08.05.21261633

**Authors:** Zoe Aitken, Eric Emerson, Anne Kavanagh

**Affiliations:** Disability and Health Unit, Centre for Health Equity, Melbourne School of Population and Global Health, The University of Melbourne; Institute for Health Research, Lancaster University

## Abstract

**Background:** COVID-19 vaccination is the cornerstone of managing Australia’s COVID-19 pandemic and the success of the vaccination program depends on high vaccination coverage. This paper examined differences in COVID-19 vaccination coverage and vaccine hesitancy for people with disability, long-term health conditions, and carers – subgroups that were prioritised in the vaccination program.

**Methods:** Using data from 2,400 Australians who participated in two waves of the Taking the Pulse of the Nation survey in April and May 2021, we described vaccination coverage and hesitancy among people with disability, severe mental health conditions, severe long-term health conditions, frequent need for assistance with everyday activities, and carers, disaggregated by age group and gender.

**Findings:** Vaccination coverage was estimated to be 8.2% in the sample overall and was similar for people with disability, those with frequent need for assistance, and carers. It was higher for people with severe long-term health conditions (13.4%) and lower for people with severe mental health conditions (4.3%). Vaccine hesitancy was high overall (35.6%) and was similarly high across the priority groups.

**Interpretation:** This study highlights the lack of a difference in vaccination coverage and vaccine hesitancy for people with disability, long-term health conditions, and carers compared to the general population. Sub-optimal vaccination coverage for people in the priority population groups leaves many people at significant risk of serious disease or death if exposed to COVID-19, particularly in light of the easing of disease-control restrictions across Australia and the emergence of new COVID-19 variants.

**Funding:** National Health and Medical Research Council

## Introduction

COVID-19 vaccination is the cornerstone of managing the COVID-19 pandemic. The success of any vaccination program depends on high vaccination coverage and acceptance. Across the world, countries implemented different strategies for the prioritisation of COVID-19 vaccination. The World Health Organization (WHO) developed guidelines for the prioritisation of vaccination among groups according to their risk of acquisition and transmission of COVID-19 (e.g., health care workers providing direct care) and risk of serious disease or death if infected by COVID-19 (e.g., older people, organ transplant recipients).^1^ WHO also recommended that the levels of community transmission in a country should also guide vaccine distribution.^1^

In Australia, the Commonwealth government has been responsible for the procurement and distribution of COVID-19 vaccines and the policy settings, although there have been some variations between States and Territories in the rollout to different groups. The Commonwealth government prioritised a range of groups in Phase 1a, which commenced on 22 February 2021 and Phase 1b which commenced on 22 March 2021 (Table 1). People with long-term health conditions including serious mental illness, people with intellectual disability and people who required assistance with activities of daily living were prioritised.^2^ These groups also included workers providing support in aged-care or to people with disability, as well as informal carers who provide unpaid support to people who needed assistance with activities of daily living.

**Table 1.** Phase 1a and 1b priority groups

| Phase 1a | Phase 1b |
| --- | --- |
| Aged care and disability care residents | People with long-term health conditions |
| Aged care and disability care staff | People with disability |
| Frontline health care workers | Carers |
| Quarantine and border workers | People aged 70 years and older |
|  | Aboriginal and Torres Strait Islander Peoples aged 56 years and older |
|  | Other health care workers |
|  | Critical and high-risk workers |

At the start of 2022, Australia had high COVID-19 vaccination coverage, ranked 15^th^ globally in terms of the proportion of the population vaccinated according to the Johns Hopkins Coronavirus Resource Centre Vaccine Tracker.^3^ By mid-February 2022, 94.1% of people aged 16 years and over had received two COVID-19 vaccine doses.^4^ However, the vaccination coverage was substantially lower in some subgroups of the population at highest risk of COVID-19 infection and morbidity, such as Aboriginal and Torres Strait Islander Peoples (76.6%) and National Disability Insurance Scheme (NDIS) participants (86.1%),^4^ demonstrating large inequalities in vaccination coverage. Therefore, despite prioritising people at highest risk, some priority groups have sub-optimal vaccination coverage which poses a threat to individual and population immunity.^5^

People with disability and long-term health conditions are at high risk of serious disease or death if infected with COVID-19 due to comorbidities, living conditions, behavioural factors and socio-economic deprivation and may have higher risk of infection and transmission because of their need for care or assistance and increased likelihood of living in congregate care settings.^6,7^ But there is a lack of data describing COVID-19 vaccination rates for people with disability and long-term health conditions because disability data are not routinely collected in Australia. Data from government and media reports from mid-2021 showed that there were significant delays in the rollout of the vaccine among people living in aged-care and disability residences and the workers who support them.^8-10^ Since 21 January 2022, data have been routinely reported on vaccination coverage for NDIS participants,^11^ demonstrating lower vaccine coverage compared to the general population, but this only represents 10% of Australians with disability. There is a lack of data on vaccination coverage for the 90% of Australians with disability who are not eligible for the NDIS.

There are several potential causes of low vaccination coverage for people with disability including physical availability of vaccines, geographic accessibility, availability of accessible information about vaccine availability and eligibility, and vaccine hesitancy.^5^ At the start of the vaccination program, there was a lack of supply – a problem that intensified after the Commonwealth government recommended that the Comirnaty (Pfizer/BioNTech) COVID-19 vaccine was used for people under 50 years on 8^th^ April 2021 and then under 60 years on 16^th^ June 2021 because of the risk of Thrombosis with Thrombocytopenia Syndrome (TTS) associated with the Vaxzevria (AstraZeneca) COVID-19 vaccine in younger age-groups. ^12,13^ Vaccine hesitancy, the delay in the acceptance or refusal of vaccines,^5,14,15^ is also a likely cause of inequalities in vaccination coverage.

Australian research has demonstrated relatively high levels of vaccine hesitancy in the population compared to similar countries.^16^ An Australian survey conducted in January 2021 before the vaccination program was implemented, found that only 44% of Australians reported that they would definitely get vaccinated, with 35% reporting they would probably get vaccinated, 8% reporting they would probably not get vaccinated, and 13% reporting they would definitely not get vaccinated.^17^ More recent data from a longitudinal survey investigating the impacts of the COVID-19 pandemic on Australian households which collected data on disability and included questions about vaccine hesitancy in five waves of the survey between December 2020 and July 2021 found that levels of vaccine hesitancy were very similar between people with and without disability, though vaccine hesitancy was higher for people with psychosocial disability.^18^

Australia experienced a surge in COVID-19 infections at the end of 2021 alongside the easing of disease-control restrictions and the emergence of the Omicron variant, which highlights the ongoing need for high COVID-19 vaccination coverage. Despite high vaccination coverage in the population overall, waning vaccine immunity and sub-optimal vaccination coverage in some subgroups of the population pose a threat to individual and population immunity.^5^ Understanding inequalities in vaccine coverage and attitudes towards vaccine hesitancy is important for tailoring vaccination programs and evidence-informed and community-engaged responses to vaccine hesitancy to different subgroups.^19,20^ It is also critical for understanding the potential impacts of easing restrictions on severe outcomes associated with COVID-19 infection, which will be distributed inequitably across different subgroups of the population.

This paper fills a gap in our knowledge about vaccination coverage and vaccine hesitancy among people with disability and long-term health conditions, and carers in Australia with the aim of identifying groups who require better targeting to improve vaccination coverage and understanding the causes of sub-optimal vaccination coverage. Using data from 2,400 Australians who participated in Taking the Pulse of the Nation survey in April and May 2021, at the start of the COVID-19 vaccination program, we describe vaccination coverage and hesitancy among people with disability, people who reported living with a severe mental health condition, a severe long-term health condition, and those requiring frequent assistance with everyday activities. We also report vaccination coverage and hesitancy for people who provide paid or unpaid care to someone in one of the priority groups.

## Methods

We used data from Taking the Pulse of the Nation (TTPN), a repeated cross-sectional survey which has been conducted every two weeks since April 2020 by the Melbourne Institute: Applied Economic & Social Research at the University of Melbourne. The survey was implemented by a market research company Oz Info as a telephone or online interview, collecting data from 1200 people at each wave to track Australians’ expectations and attitudes towards the COVID-19 pandemic. The sampling frame was constructed by Oz Info from a diverse set of continuously updated proprietary databases. A stratified sampling design was employed, dividing the population into strata defined by selected characteristics, which were sampled separately to ensure representativeness of the underlying population of interest.^21^ The strata were defined according to state of residence (all six states and the Australian Capital Territory), location within each state (Greater Capital Area; other), gender (men; women) and age group (18-24; 25-34; 35-44; 45-54; 55-64; 65-75; older than 75 years), with weights constructed to ensure the sample in each wave was representative of the underlying population using ABS estimated resident population projections based on the 2016 Census.^22^ The TTPN survey was approved by the Human Research Ethics Committee at the University of Melbourne (2021-14006-14669-1). The TTPN Survey data are available on application to the Melbourne Institute: Applied Economic & Social Research at the University of Melbourne.

This analysis used data from two waves of the survey conducted between 19 and 25 April and between 10 and 14 May 2021 in which additional questions were included about disability, long-term health conditions and caring responsibilities. At this stage of the pandemic, people over the age of 50 years and people in high priority groups (and their carers) were eligible to receive the vaccine (Table 1). In these two waves of the survey, a question was included about presence of disability, defined as “a long-term health condition, impairment or disability that restricts you in your everyday activities and has lasted (or is likely to last) for 6 months or more”. Additional questions were included to identify whether people had a severe mental health condition, a severe long-term health condition, or required frequent assistance with everyday activities (e.g., eating, dressing, mobility), to align with the vaccine priority group eligibility criteria. A question was also included to identify people who provided care, help or assistance, either paid or unpaid, to people in the high priority groups. Survey participants could tick multiple responses to these questions. For all questions, responses “don’t know” were recoded to missing.

Information on vaccination and vaccine hesitancy was recorded in the survey. Participants were asked if they were “willing to have the COVID-19 vaccine” with possible responses listed as “I have had it already”, “yes”, “no”, and “don’t know”. People were identified as being vaccinated if they responded that they had had the vaccine. Therefore, vaccination coverage represents people who have had at least one dose of the vaccine. People were identified as being vaccine hesitant if they responded “no” or “don’t know”, including both vaccine refusers and those who may choose to delay, in accordance with the definition derived by the SAGE Working Group on Vaccine Hesitancy.^23^ People were identified as not vaccine hesitant if they responded “yes” or were already vaccinated. We included people who were vaccinated in the non-vaccine hesitant group to ensure valid comparisons of vaccine hesitancy between subgroups of the population, independent of the proportion of people who had been vaccinated.

Population subgroups of interest included age groups (18-64 years; 65 years and above) and gender (men; women).

We estimated the proportion of the sample who were vaccinated and who were vaccine hesitant, for the whole sample (both waves combined) and for exposure groups of interest (people with disability, severe mental health conditions, severe long-term health conditions, people requiring frequent assistance with everyday activities, and people who were paid or unpaid carers). We disaggregated the results by age group and gender. Results were not presented separately for each wave of the survey due to sample size limitations. The analyses used survey weights, constructed using the most recent Australian Bureau of Statistics estimated resident population projections based on the 2016 Census, stratified by gender, age and location to be representative of the Australian population. Analyses were conducted in Stata/SE 14.2 using the survey commands to account for the survey design characteristics.

## Results

The two waves of the survey included 2,400 participants, of which 8.2% were vaccinated and 35.6% were vaccine hesitant (Table 2), comprising 19.1% who reported being unwilling to get the vaccine and 16.5% who were unsure (data not shown).

**Table 2.** Characteristics of the sample (n=2,400)

|  | n | % |
| --- | --- | --- |
| Vaccinated |  |  |
| Yes | 177 | 8.2 |
| No | 2,223 | 91.8 |
| Vaccine hesitancy |  |  |
| Yes | 854 | 35.6 |
| No | 1,546 | 64.4 |
| Disability |  |  |
| No | 1,623 | 69.9 |
| Yes | 709 | 27.5 |
| Missing | 68 | 2.6 |
| Severe mental health condition |  |  |
| No | 2,118 | 89.2 |
| Yes | 282 | 10.8 |
| Missing | 0 | 0 |
| Severe long term health condition |  |  |
| No | 1,913 | 80.1 |
| Yes | 487 | 19.9 |
| Missing | 0 | 0 |
| Frequent need for assistance |  |  |
| No | 2,258 | 94.4 |
| Yes | 142 | 5.6 |
| Missing | 0 | 0 |
| Carer |  |  |
| No | 1,700 | 71.5 |
| Yes | 655 | 26.4 |
| Missing | 45 | 2.1 |
| Age group |  |  |
| 18-64 years | 1,942 | 79.8 |
| 65 years and older | 458 | 20.2 |
| Missing | 0 | 0 |
| Gender |  |  |
| Men | 1,200 | 49.0 |
| Women | 1,200 | 51.0 |
| Missing | 0 | 0 |

Of the sample, 27.5% of participants had a disability, 10.8% had a severe mental health condition, 19.9% had a severe long term health condition, 5.6% had frequent need for assistance, and 26.4% were paid or unpaid carers. In terms of demographics, 20.2% of the sample were aged 65 years and older, and 51% were women.

### Vaccination coverage

Compared to the sample overall, the proportion of people who had been vaccinated was higher for people with severe long term health conditions (13.4%), similar for those with disability (8.9%), people in need of frequent assistance with everyday activities (7.1%), and carers (8.9%), and lower for people with severe mental health conditions (4.3%, Table 3). Vaccination coverage was similar for men (9.0%) and women (7.4%), and the patterns across the priority groups were broadly consistent with the sample overall when disaggregated by gender. Vaccination coverage was higher for people aged 65 years and older (30.5%) compared to those aged 18 to 64 years who had very low vaccination coverage (2.6%), even for people in the vaccine priority groups, ranging from 1.2% to 4.2%.

**Table 3.** Proportion of the study population who had received at least one dose of COVID-19 vaccine, by priority group, age and gender

|  | Overall | Disability | Severe mental health condition | Severe long-term health condition | Frequent need for assistance | Carer |
| --- | --- | --- | --- | --- | --- | --- |
|  | % (95% CI) | % (95% CI) | % (95% CI) | % (95% CI) | % (95% CI) | % (95% CI) |
| Whole sample | 8.2 (6.6, 9.8) | 8.9 (6.8, 11.6) | 4.3 (2.5, 7.5) | 13.4 (9.9, 17.8) | 7.1 (3.6, 13.4) | 8.9 (6.4, 12.2) |
| Age |  |  |  |  |  |  |
| 18-64 | 2.6 (1.8, 3.6) | 3.3 (2.0, 5.5) | 2.3 (1.2, 4.5) | 4.2 (2.4, 7.4) | 1.2 (0.4, 3.8) | 4.0 (2.2, 7.1) |
| 65+ | 30.5 (24.9, 36.8) | 27.5 (20.1, 36.4) | 42.6 (19.1, 70.0) | 32.7 (23.5, 43.4) | 26.4 (12.1, 48.2) | 26.2 (18.0, 36.5) |
| Gender |  |  |  |  |  |  |
| Men | 9.0 (6.9, 11.8) | 10.7 (7.5, 15.0) | 3.0 (1.2, 7.1) | 14.0 (9.9, 19.5) | 7.8 (3.2, 17.8) | 8.4 (5.6, 12.4) |
| Women | 7.4 (5.6, 9.7) | 7.3 (4.8, 11.0) | 5.9 (2.8, 11.7) | 12.8 (7.9, 20.2) | 6.1 (2.2, 16.0) | 9.3 (5.8, 14.5) |

### Vaccine hesitancy

Overall vaccine hesitancy was high, with 35.6% of the sample estimated to be hesitant to receive the vaccine (Table 4). Vaccine hesitancy was found to be similarly high across the priority groups, ranging from 24.2% for people with frequent need for assistance to 36.1% for people with severe mental health conditions. Vaccine hesitancy was higher for people aged 18 to 64 years (39.7%) compared to those aged 65 years and older (19.3%) and patterns within each age group were similar across the priority groups. There was a slight indication than vaccine hesitancy was higher for women than men (39.9% versus 31.1%).

**Table 4.** Proportion of the study population who were vaccine hesitant, by priority group, age and gender

|  | Overall<br>% (95% CI) | Disability<br>% (95% CI) | Severe mental<br>health condition<br>% (95% CI) | Severe long-term<br>health condition<br>% (95% CI) | Frequent need for<br>assistance<br>% (95% CI) | Carer<br>% (95% CI) |
| --- | --- | --- | --- | --- | --- | --- |
| Whole sample | 35.6 (33.2, 38.0) | 30.3 (26.3, 34.7) | 36.1 (29.4, 43.5) | 27.7 (23.0, 33.0) | 24.2 (16.6, 33.8) | 30.7 (26.5, 35.2) |
| Age |  |  |  |  |  |  |
| 18-64 | 39.7 (37.0, 42.4) | 34.7 (30.0, 39.8) | 37.4 (30.4, 45.0) | 32.9 (26.9, 39.6) | 27.5 (18.5, 38.9) | 34.3 (29.4, 39.6) |
| 65+ | 19.3 (15.3, 24.1) | 15.7 (10.6, 22.8) | 12.3 (2.9, 39.1) | 16.7 (11.0, 24.6) | 13.4 (5.0, 31.2) | 18.0 (11.8, 26.4) |
| Gender |  |  |  |  |  |  |
| Men | 31.1 (27.9, 34.5) | 28.1 (22.6, 34.4) | 30.3 (21.3, 41.0) | 27.1 (20.4, 35.0) | 25.6 (14.9, 40.3) | 22.8 (17.5, 29.1) |
| Women | 39.9 (36.5, 43.4) | 32.3 (26.7, 38.6) | 42.8 (33.1, 53.1) | 28.2 (21.8, 35.6) | 22.5 (13.6, 34.9) | 36.3 (30.3, 42.8) |

## Discussion

The findings of this study suggest that vaccination coverage was similar between people in the overall sample and the majority of the priority groups examined in this study, including people with disability, people with frequent need for assistance and people who were carers. There was some evidence that vaccination coverage was higher for people with severe long-term health conditions and lower for people with severe mental health conditions. Vaccination coverage was strikingly low for younger people in all priority groups. Given that people in these priority groups were eligible to receive the vaccine at the time of the survey, the lack of difference in vaccination coverage between young people in the priority groups and the overall sample is surprising and of concern.

Despite people with disability being prioritised for vaccination, the Royal Commission hearing in May 2021 highlighted the low rates of vaccination for people with disability.^24^ The results of this study were consistent with the findings of the Royal Commission, indicating low vaccination coverage and little evidence of a difference in vaccination coverage for people with disability and long term health conditions compared to the overall sample.

Our study found that vaccination coverage was higher for people aged 65 years and older compared to those aged 18 to 64 years. Older adults aged 70 years and older became eligible for vaccination in phase 1b, which explains the higher coverage among older people in the sample, driven by age-based eligibility. This suggests the presence of additional barriers to achieving high vaccination coverage for people with disability and long-term health conditions for whom eligibility for vaccination did not translate into higher vaccination coverage.

We found evidence of high levels of vaccine hesitancy overall with more than one in three people in the sample reporting that they were hesitant to receive a COVID-19 vaccine. There were high levels of vaccine hesitancy for people in the priority groups, particularly for young people, despite their elevated risk of serious illness or death if they contracted COVID-19. It is important to note that data for the first of the two waves of the survey were collected just after the change in the Commonwealth government recommendation that people under 50 years were to receive the Cominraty (Pfizer/BioNTech) vaccine rather than the Vaxzevria (AstraZeneca) vaccine because of the risk of TTS. This announcement may account, in part, for the elevated vaccine hesitancy reported in this study, particularly for young people. Indeed, vaccine hesitancy in the first wave for which data were available on priority groups was estimated to be 39% overall, notably higher than previous and subsequent waves of the TTPN survey where vaccine hesitancy was 32% on average, ranging from 29% to 35%. The estimates of vaccine hesitancy are consistent in magnitude with statistics published by the government which suggest that vaccine hesitancy in Australian adults peaked in April and May 2021 and then gradually declined.^25^ Given Australia’s current high vaccination rates, vaccine hesitancy at the start of the vaccination program has not translated into vaccine refusal for a large proportion of the population. However, data are not currently available on the trajectory of vaccination uptake (and vaccine hesitancy) for people with disability and long-term health conditions, which may be different to the overall population.

Given the lack of difference in vaccine hesitancy between people with disability and long-term health conditions and the overall sample, it is unlikely that the low vaccination coverage for people with disability and long-term health conditions is due to increased vaccine hesitancy. Reports from disability organisations and media have suggested that people with disability experienced multiple barriers, including problems securing appointments, lack of accessible information about eligibility and availability of vaccines, and finding accessible vaccination clinics.^26,27^ The lack of research examining reasons for low vaccination coverage is an important evidence gap, and important to address for COVID-19 vaccination but also for other vaccines.

Our study had a number of strengths. This is the first study to examine vaccination coverage and hesitancy for people with disability and long-term health conditions in Australia, using definitions aligning closely with the vaccine priority groups. The sample was large enough to identify people in all the priority groups of interest, including analyses disaggregated by age group and gender. There were also limitations. We did not examine all vaccine priority groups. There was insufficient data to examine occupational vaccine priority groups such as healthcare workers, though it would be valuable to gain insights into vaccination coverage and hesitancy in these groups who were also eligible to receive the vaccine. We only examined two broad age groups because there were too few younger people in the priority groups to disaggregate the age groups further. As a result, we could not examine people aged younger than 50 years separately, for whom there were different recommendations for use of the Cominraty (Pfizer/BioNTech) vaccine. Further, due to the age categories used in the survey, it was not possible to identify people aged 70 years and above to align with age-based eligibility. We did not examine differences by other socio-demographic characteristics including ethnicity, Aboriginal and Torres Strait Islander peoples, or cultural and linguistic diversity because the survey did not collect data on these characteristics. Given the evidence about differences in vaccine hesitancy by ethnicity in the United Kingdom and the Unites States^28^ but not found in New Zealand^29^ it is important to examine differences in vaccine hesitancy according to ethnicity for people in priority groups in future research. Participation rates could not be estimated in this study, therefore it was not possible to assess the impact of potential selection bias from non-participation. Due to its relatively small sample, the survey is unlikely to be representative of the Australian population, though the sample weights make the results more representative of the population. The vaccine coverage was estimated to be 8.2% overall, which is broadly consistent, though perhaps slightly lower, than population estimates of vaccination coverage at similar time points, estimated to be 7.5% on 25^th^ April 2021 and 12.1% on 16^th^ May 2021.^30^

## Conclusions

This study highlights the lack of a difference in vaccination coverage in the first few months of Australia’s COVID-19 vaccination program between people with disability and long-term health conditions who were prioritised in the COVID-19 vaccination program and the general population. The results are concerning because low vaccination coverage for people in the priority population groups leaves many people at significant risk of serious disease or death if exposed to COVID-19, particularly in light of the easing of disease-control restrictions across Australia once population vaccination targets were achieved and the emergence of new COVID-19 variants.

Recent data are needed to analyse current inequalities in vaccination coverage between people with disability and long-term health conditions compared to the overall population statistics to contrast the results from the start of the vaccination program with the current situation in which Australia has reached high vaccination coverage in the population overall. Furthermore, data should be routinely collected to report on vaccination coverage for people with disability and long-term health conditions.

Vaccine hesitancy was found to be similar between people with disability and long-term health conditions compared to the overall sample, suggesting that low vaccination coverage is likely to be caused by barriers to accessing vaccination rather than high vaccine hesitancy. There remains a lack of understanding of the barriers faced by people in these priority groups. Further research is needed to understand the barriers to accessing COVID-19 vaccination experienced by Australians with disability and long-term health condition, to generate the information that is needed to devise effective strategies to improve uptake by ensuring these populations have easy access to COVID-19 vaccines such as accessible vaccination hubs, in-reach into workplaces and homes, and co-designed communication strategies.

## Data Availability

Researchers can access the Taking the Pulse of the Nation Survey for research and replication purposes. To gain access, please contact Dr. Nicolas Salamanca at.

## Authors’ contributions

ZA, AK and EE conceived the study and designed the questions for the TTPN Survey. ZA analysed the data and results were reviewed and interpreted by ZA, AK and EE. ZA and AK wrote the manuscript, which was reviewed by EE.

## Ethics committee approval

The Taking the Pulse of the Nation Survey was approved by the Human Research Ethics Committee at the University of Melbourne (2021-14006-14669-1).

